# Systems strengthening approach during antenatal care improves maternal nutrition and reduces childhood stunting in West Bengal, India

**DOI:** 10.1101/2022.10.14.22281107

**Authors:** Kayur Mehta, Sreeparna Ghosh Mukherjee, Ipsita Bhattacharjee, Kassandra Fate, Shivani Kachwaha, Aastha Kant, Meghendra Banerjee, Anita Shet

## Abstract

**Background:** Early childhood growth failure including stunting is associated with suboptimal health and cognitive development outcomes. Despite progress, the prevalence of childhood stunting in India remains amongst the highest globally.

**Objective:** We aimed to evaluate the impact of a systems strengthening interventional package, including body-mass-index measurement at pregnancy registration, monthly weight monitoring, on-the-spot supplementary nutrition, iron-folic acid supplementation, and targeted dietary counselling provided to women during their antenatal care on childhood stunting.

**Methods:** This is a prospective follow-up comparison study. Women from three districts in West Bengal, India in their first trimester of pregnancy between May 2018 and May 2019 were enrolled into the study. Pregnancy, birth and infant characteristics were collected, and anthropometric indices measured. The relative risk of stunting in children in intervention and comparison groups were compared using generalized linear model to adjust for clustering effect.

**Results:** A total of 809 mother-child dyads (406 intervention; 403 comparison) were followed between May 2018 and May 2021. The median age of women in the intervention and comparison group was 23 (IQR 20-25) and 25 (IQR 24-27) years respectively. Median gestational weight gain was higher amongst women in the intervention group (9 vs. 8 kilograms, p=0.04). Low-birth-weight prevalence was 29.3% (119/406) and 32.0% (129/403) in the intervention and comparison group respectively. At 12-35 months of age, children born to women in the intervention group had significantly reduced risk of stunting (RR=0.58, 95% CI 0.45-0.75, p<0.001). The odds of stunting amongst children born with low birthweight to women in the comparison group were statistically significant [OR 2.44 (1.44-4.14)], unlike those amongst children born to women in the intervention group [1.19 (0.58-2.46)].

**Conclusions:** These results indicate that strengthening of routine antenatal care including targeted nutritional counselling to expectant mothers can have distal beneficial effects on childhood stunting beyond the immediate post-natal period.

**Teaser Text:** This article describes the impact of a systems strengthening approach during antenatal care that improved maternal nutrition and reduced childhood stunting in West Bengal, India.

## Background

The past decade has witnessed a renewed focus on childhood undernutrition, particularly on stunting. The inclusion of stunting targets in the Sustainable Development Goals has further enthused efforts to work towards the World Health Assembly’s 2012 nutrition target of 40% reduction in the number of children under 5 years (U5) who are stunted by 2025(1, 2). However, global progress on stunting reduction among U5 children has been slow; as per the 2021 UNICEF/WHO/World Bank Group Joint Child Malnutrition estimates, approximately 149 million children under-five (22%) were stunted, compared to 144 million U5 children (21.3%) in 2019(3).

India carries one of the largest global burdens of maternal and child undernutrition, approximately one third of the world’s total population of stunted children under 5 years of age who are stunted reside in India(4). As per National Family Health Survey-5 (NFHS-5) estimates (2019-2021), the fraction of U5 children in India who are stunted is 35.5%, a value far above the global proportion of 27%, and distant from the targets set under the Sustainable Development Goals. In the Indian state of West Bengal, according to the National Family Health Survey-5 (NFHS-5, 2019-2021) estimates, 34% children under the age of 5 years are stunted(4), a proportion that has increased from the previous survey conducted five years earlier (NFHS-4, 2015-2016)(5).

Stunting is the result of poor nutrition in-utero and early childhood and is associated with numerous short-term and long-term consequences, including increased childhood morbidity and mortality(6), delayed growth and motor development(7), and long-term educational and economic associations, including poor scholastic performance, increased absences from work and lower productivity(8). Maternal nutrition plays a vital role in fetal growth, infant health and survival as well as long-term child health and development(9). Maternal undernutrition during pregnancy is a major determinant of poor fetal growth and child stunting(10).

In order to examine the effect of maternal interventions on child growth, we conducted an extension of a nutritional intervention project for expectant mothers that was initiated in 2018-19 in selected districts of West Bengal that studied pregnancy and birth outcomes(11). The extension study reported here was conducted in 2021-2022 to assess the prevalence of stunting amongst children born to women in the intervention and comparison groups.

## Methods

### Study design and sample population

This was a prospective follow-up comparison study. The original maternal and child health and nutrition project consisting of intervention and comparison groups was conducted in between 2018-2021 in selected blocks of three districts of the state of West Bengal: Nagrakata block from Jalpaiguri district; Suti-I block from Murshidabad district; and Falta block from South 24 Parganas district in West Bengal, India. For the current comparison study, mother-child dyads from the same districts were followed up from May 2018 until May 2021. Women in their first trimester of pregnancy in May 2018 and May 2019 at the original intervention Integrated Child Development Services (ICDS) centers in these districts were included in the current intervention group (n=485). The number in the intervention group available for follow-up was 406; exclusions were due to migration (n=34), lost to follow up (n=21), abortion or miscarriage (n=10), stillbirths (n=7) and child deaths (n=7) (Figure 1). Age matched women from the same geographic areas who were pregnant during the same time frame and did not receive the interventions, and were currently available for follow-up constituted the comparison group (n=403), which was chosen at the time of prospective data collection for this study.

**Fig. 1:**
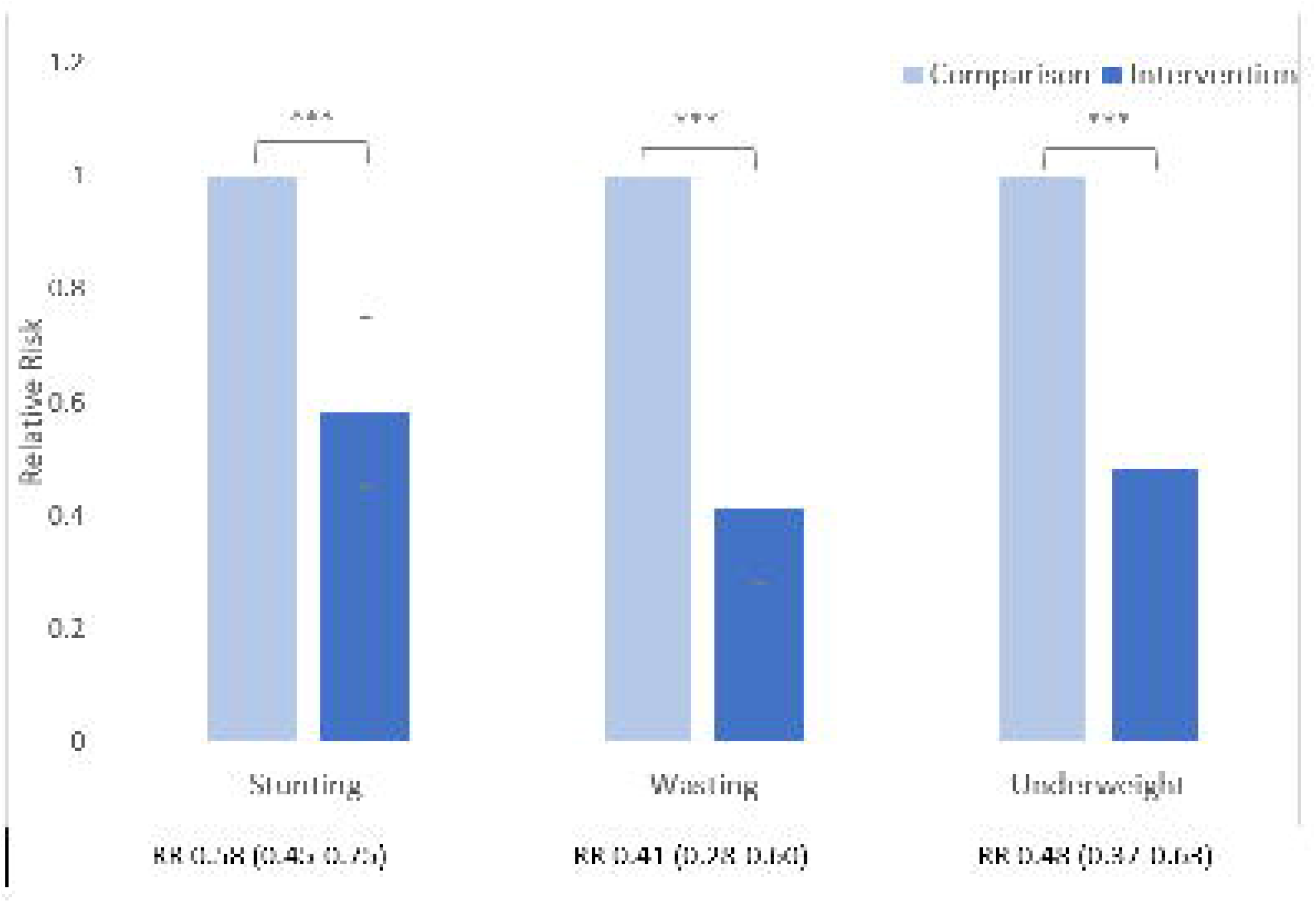
Impact of the intervention on nutritional status in children aged between 12-35 months Legend: Note: *p<0.05, **p<0.01, ***p<0.001 denotes differences in RR between intervention and comparison groups

### Intervention

In the original study, pregnant women in the intervention group received a bundle of interventions that were contained within national guidelines(12), that were strengthened with added support in implementation from the research team. Strengthening interventions, which bolstered those in the national guidelines included the following: (i) baseline maternal BMI measurement and monthly weight monitoring, (ii) ensuring daily on-the-spot supplementary nutrition for expectant mothers at ICDS centers, (iii) ensuring antenatal iron and folic acid supplementation, and (iv) providing targeted dietary counselling based on nutritional status (BMI) as well as joint counselling of family members to ensure support for the expectant mother and sharing of household work burden and (v) identification and follow up of pregnant women at “nutritional risk” (A pregnancy was considered ‘at nutritional risk’ when at least one of the following indicators were present: a) BMI (taken <20 weeks gestation) identified woman as severely thin, thin, overweight or obese b) Age of pregnancy (below 20 and above 35 years); c) Body weight at the time of registration (40 kg or less); d) Height (less than 145 cm); e) Anemia (severe anemia: Hemoglobin less than 7 g/dl, moderate anaemia: 7-10.9 g/dl); f) Inappropriate gestational weight gain (GWG) (<1 kg /month or >3 kg /month from second trimester onwards(13). These interventions were also strengthened by ensuring increased home contact amongst identified women to ensure timely uptake of antenatal services, hemoglobin testing and treatment of anemia, ensuring ultrasonography after the second trimester and ensuring a fourth antenatal visit in the ninth month of pregnancy [all included in the national guidelines(12)].

### Outcomes

In the current study, follow-up was conducted in 2021-2022 when the offspring of interventions and comparison women were 12-35 months of age. The primary outcome was stunting, defined as length-for-age <-2 standard deviations (SD) from the median of the World Health Organization (WHO) child growth standards among 12–35-month-old children of women in the intervention group and comparison groups. Other outcomes among these children were wasting and underweight, defined respectively as weight-for-length <-2 SD and weight-for-age <-2 SD from the median of the WHO child growth standards.

### Data collection

In the original study, data were collected by community health workers during house visits to pregnant women in the intervention group. Demographic details were collected during interviews, and data regarding pregnancy, birth and infant characteristics were abstracted from Maternal and Child Protection Cards. Details including maternal age, parity, birth interval, BMI at first antenatal visit, baseline maternal hemoglobin status (obtained from hemoglobin measurements during the first antenatal visit), maternal education status, number of antenatal visits, gestational weight gain, maternal hemoglobin at the third antenatal visit and mode of delivery were collected. Child details including the gestational age at birth, birth weight, and breastfeeding practices were obtained. In this follow-up study, similar data were collected form the women who constituted the comparison group. Standard anthropometric equipment was used to measure the weight and height of the pregnant women, and weight and length or height of the children.

### Statistical analysis

Descriptive analyses were performed to assess the distribution of variables between the intervention and comparison groups. Proportions and medians (with interquartile ranges) are presented for baseline variables. Chi-square tests were used to compare variables between intervention and comparison groups. Anthropometric measures together with the age and sex of the children were used to calculate the weight-for-age (underweight), height for-age (stunting) and weight-for-height (wasting) z-scores according to the WHO growth standards. Relative risk of stunting, wasting, and underweight in children were compared between intervention and comparison groups using generalized linear model (GLM) to adjust for clustering effect. The log (link) function was used for binary outcomes. Multivariable logistic regression analyses were conducted to identify pregnancy and newborn related factors associated with stunting in the intervention and comparison groups. All analyses were performed using STATA version 17.

### Ethical considerations

Written informed consent was obtained from parents or caregivers at the time of child enrolment in the current study. Ethics approval was obtained from the Johns Hopkins Bloomberg School of Public Health Institutional Review Board (No. 18366) and the Institutional Ethics Committee at the Child in Need Institute, Kolkata, India (No. 01/2021-22).

## Results

### Maternal and Newborn Characteristics

#### Antenatal factors

A total of 809 women with living offspring (406 in the intervention group; 403 in the comparison group) were included in the study. The median age of women was 23 years (IQR 20-25) and 25 years (IQR 24-27) in the intervention and comparison groups respectively. Fifty-three percent (217/406) women in the intervention group and 44.4% (179/403) women in the comparison group were primiparous. At the time of pregnancy registration, 26.1% (106/406) women in the intervention group and 31.9% (128/403) women in the comparison group were underweight, and 11.5% (47/406) of the women in the intervention group and 10.9% (44/403) of the women in the comparison group were overweight. Baseline hemoglobin levels were normal (≥ 11g/dl) among 46.1% (187/406) in the intervention group and 44.1% (176/403) in the comparison group. Four or more antenatal visits were recorded for 83.9% (341/406) women in the intervention group and 96.3% (388/403) women in the comparison group (Table 1).

**Table 1.**
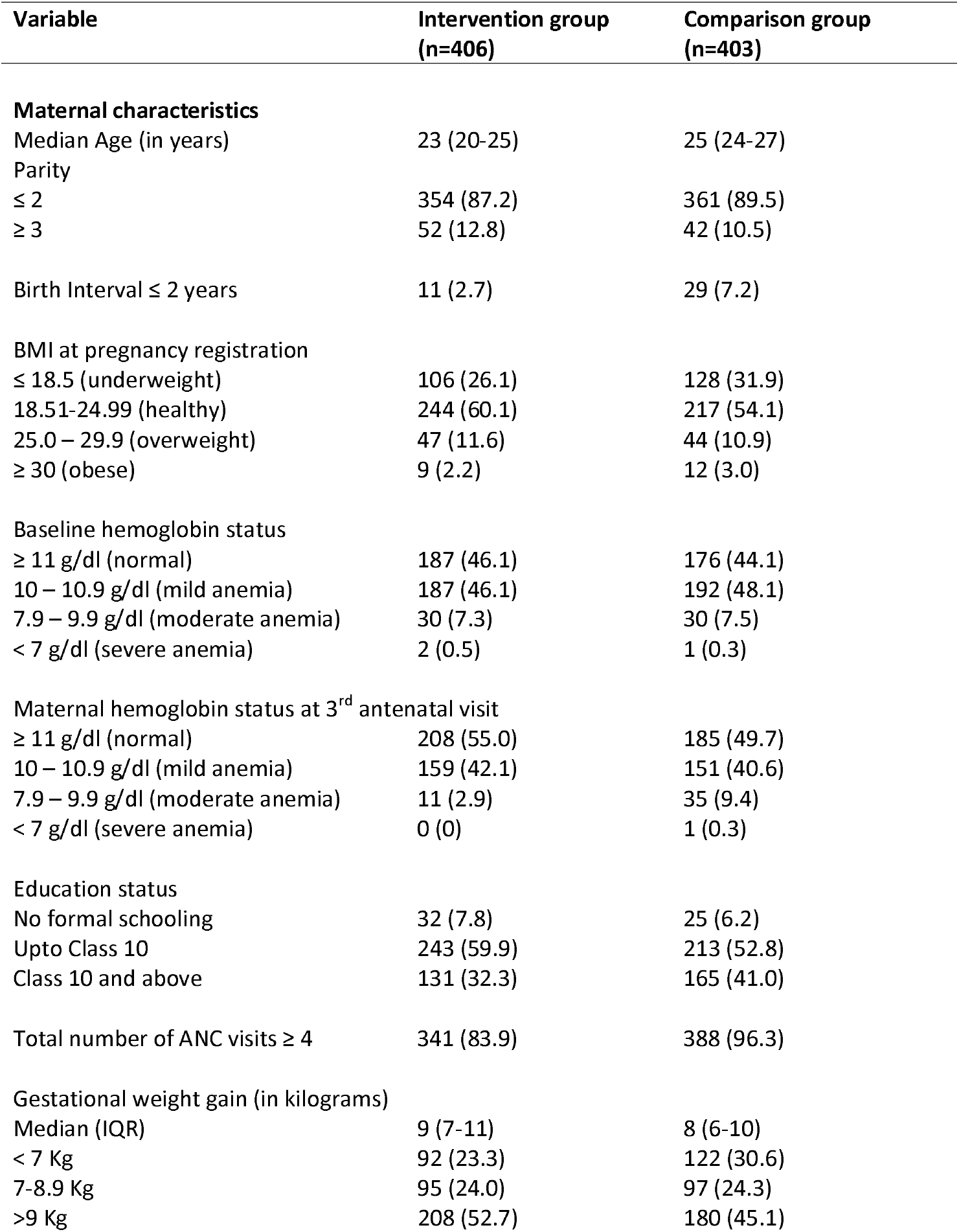
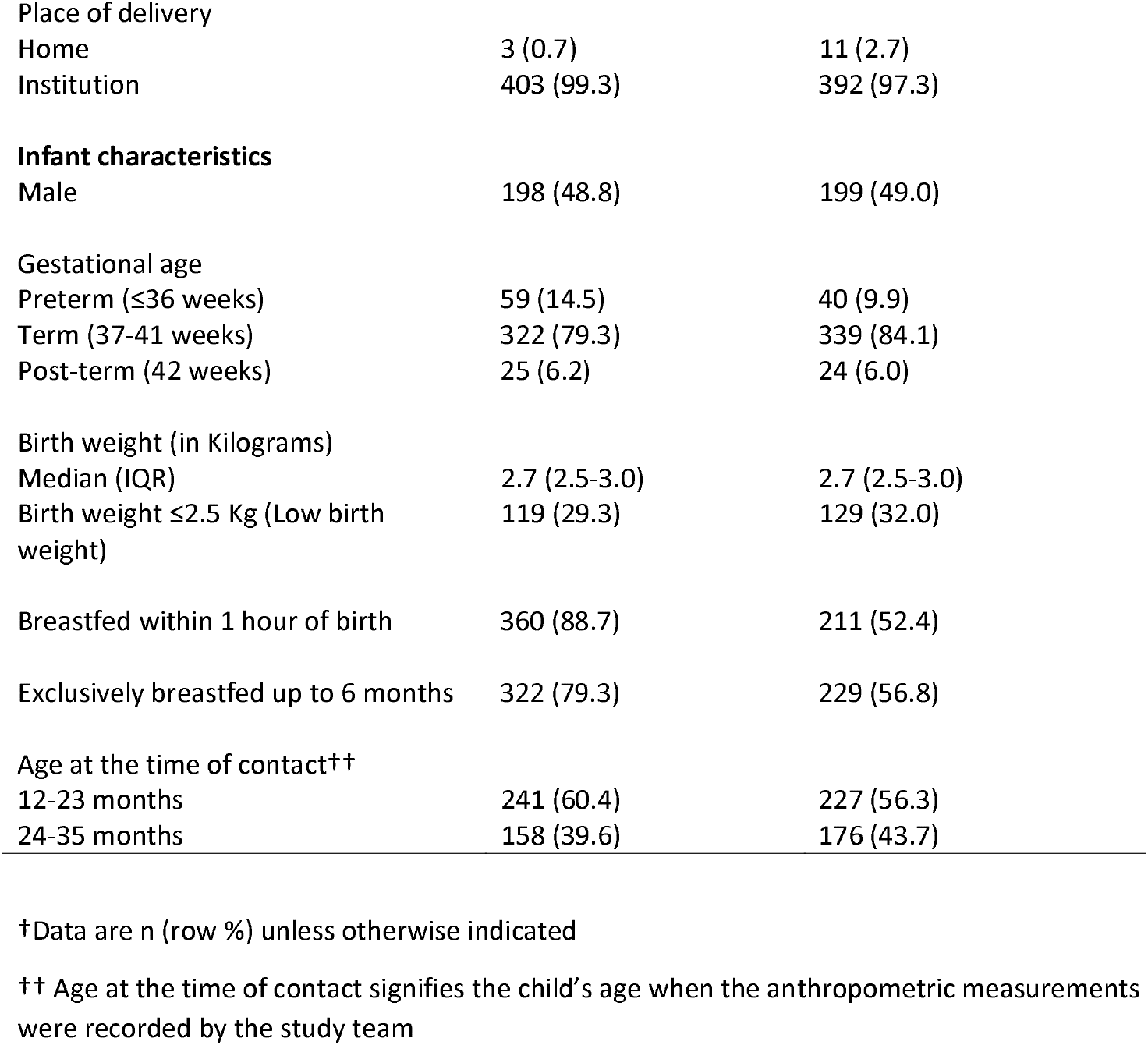
Characteristics of mothers and children (n=809) †

| Variable | Intervention group<br>(n=406) | Comparison group<br>(n=403) |
| --- | --- | --- |
| **Maternal characteristics** |  |  |
| Median Age (in years) | 23 (20-25) | 25 (24-27) |
| Parity |  |  |
| ≤ 2 | 354 (87.2) | 361 (89.5) |
| ≥ 3 | 52 (12.8) | 42 (10.5) |
| Birth Interval ≤ 2 years | 11 (2.7) | 29 (7.2) |
| BMI at pregnancy registration |  |  |
| ≤ 18.5 (underweight) | 106 (26.1) | 128 (31.9) |
| 18.51-24.99 (healthy) | 244 (60.1) | 217 (54.1) |
| 25.0 – 29.9 (overweight) | 47 (11.6) | 44 (10.9) |
| ≥ 30 (obese) | 9 (2.2) | 12 (3.0) |
| Baseline hemoglobin status |  |  |
| ≥ 11 g/dl (normal) | 187 (46.1) | 176 (44.1) |
| 10 – 10.9 g/dl (mild anemia) | 187 (46.1) | 192 (48.1) |
| 7.9 – 9.9 g/dl (moderate anemia) | 30 (7.3) | 30 (7.5) |
| < 7 g/dl (severe anemia) | 2 (0.5) | 1 (0.3) |
| Maternal hemoglobin status at 3<sup>rd</sup> antenatal visit |  |  |
| ≥ 11 g/dl (normal) | 208 (55.0) | 185 (49.7) |
| 10 – 10.9 g/dl (mild anemia) | 159 (42.1) | 151 (40.6) |
| 7.9 – 9.9 g/dl (moderate anemia) | 11 (2.9) | 35 (9.4) |
| < 7 g/dl (severe anemia) | 0 (0) | 1 (0.3) |
| Education status |  |  |
| No formal schooling | 32 (7.8) | 25 (6.2) |
| Upto Class 10 | 243 (59.9) | 213 (52.8) |
| Class 10 and above | 131 (32.3) | 165 (41.0) |
| Total number of ANC visits ≥ 4 | 341 (83.9) | 388 (96.3) |
| Gestational weight gain (in kilograms) |  |  |
| Median (IQR) | 9 (7-11) | 8 (6-10) |
| < 7 Kg | 92 (23.3) | 122 (30.6) |
| 7-8.9 Kg | 95 (24.0) | 97 (24.3) |
| >9 Kg | 208 (52.7) | 180 (45.1) |
| Place of delivery |  |  |
| Home | 3 (0.7) | 11 (2.7) |
| Institution | 403 (99.3) | 392 (97.3) |
| **Infant characteristics** |  |  |
| Male | 198 (48.8) | 199 (49.0) |
| Gestational age |  |  |
| Preterm ($\leq 36$ weeks) | 59 (14.5) | 40 (9.9) |
| Term (37-41 weeks) | 322 (79.3) | 339 (84.1) |
| Post-term (42 weeks) | 25 (6.2) | 24 (6.0) |
| Birth weight (in Kilograms) |  |  |
| Median (IQR) | 2.7 (2.5-3.0) | 2.7 (2.5-3.0) |
| Birth weight $\leq 2.5$ Kg (Low birth weight) | 119 (29.3) | 129 (32.0) |
| Breastfed within 1 hour of birth | 360 (88.7) | 211 (52.4) |
| Exclusively breastfed up to 6 months | 322 (79.3) | 229 (56.8) |
| Age at the time of contact<sup>††</sup> |  |  |
| 12-23 months | 241 (60.4) | 227 (56.3) |
| 24-35 months | 158 (39.6) | 176 (43.7) |

<sup>†</sup>Data are n (row %) unless otherwise indicated
<sup>††</sup> Age at the time of contact signifies the child's age when the anthropometric measurements were recorded by the study team

#### Pregnancy outcomes

Median gestational weight gain recorded was greater amongst women in the intervention group (9 kilograms) compared to those in the comparison group (8 kilograms, p=0.04). A greater proportion of women in the intervention group (208/406, 55.0%) compared to those in the comparison group (185/403, 49.7%) recorded normal hemoglobin levels at the 3rd antenatal visit (p=0.09) (Table 1).

#### Newborn outcomes

Births in a healthcare facility were recorded for 98.2% (795/809) children; birth at full-term was recorded for 79.3% (322/406) children in the intervention group and 84.1% (339/403) of the children in the comparison group. Low birth weight prevalence was 29.3% (119/406) in the intervention group and 32.0% (129/403) in the comparison group. Breastfeeding was commenced within 1 hour of birth among 88.7 % (360/406) women in the intervention group versus 52.4% (211/403) women in the comparison group (p<0.001). Exclusive breastfeeding up to 6 months of age was reported among 79.3% (322/406) women in the intervention group compared to 56.8% (229/403) women in the comparison group (p<0.001) (Table 1).

#### Child growth outcomes

The prevalence of stunting was 17.8% (72/399) and 30.8% (124/403) among children aged between 12-35 months in the intervention and comparison groups respectively (RR = 0.58, 95% CI: 0.45-0.75, p = <0.001) (Figure 1). The reduced RR for stunting was consistent among age subgroups 12-23 months and 24-35 months. (Supplementary figures 1 and 2).

The prevalence of wasting amongst children aged between 12-35 months in the intervention group was 8.0% (32/399) compared to 19.7% (79/401) amongst children in the comparison group (RR = 0.41, 95% CI 0.28-0.60, p <0.001). The prevalence of underweight amongst children aged between 12-35 months in the intervention group was 16.0% (64/399) compared to 33.2% (134/403) amongst children in the comparison group (RR = 0.48, 95% CI 0.37-0.63, p <0.001). Subgroup analysis by age also showed reduced risks of both these outcomes amongst children born to women in the intervention group (Supplementary figures 1 and 2).

#### Pregnancy and newborn related factors associated with childhood stunting in the intervention and comparison groups

Within the intervention group, the odds of stunting at 12-35 months among children born with low birth weight were not significantly different (OR 1.16, 95% CI 0.55-2.45) from those with normal birth weight; whereas within the comparison group, children born with low birth weight had significant odds of being stunted (OR 2.54, 95% CI 1.49-4.34) compared to children born with normal birth weight within this group (Figure 1). The odds of stunting amongst children in the intervention and comparison groups related to other pregnancy related factors such as the number of antenatal visits, maternal anemia at the third antenatal visit and gestational weight gain were not statistically different between the two groups (Figure 2).

**Fig. 2:**
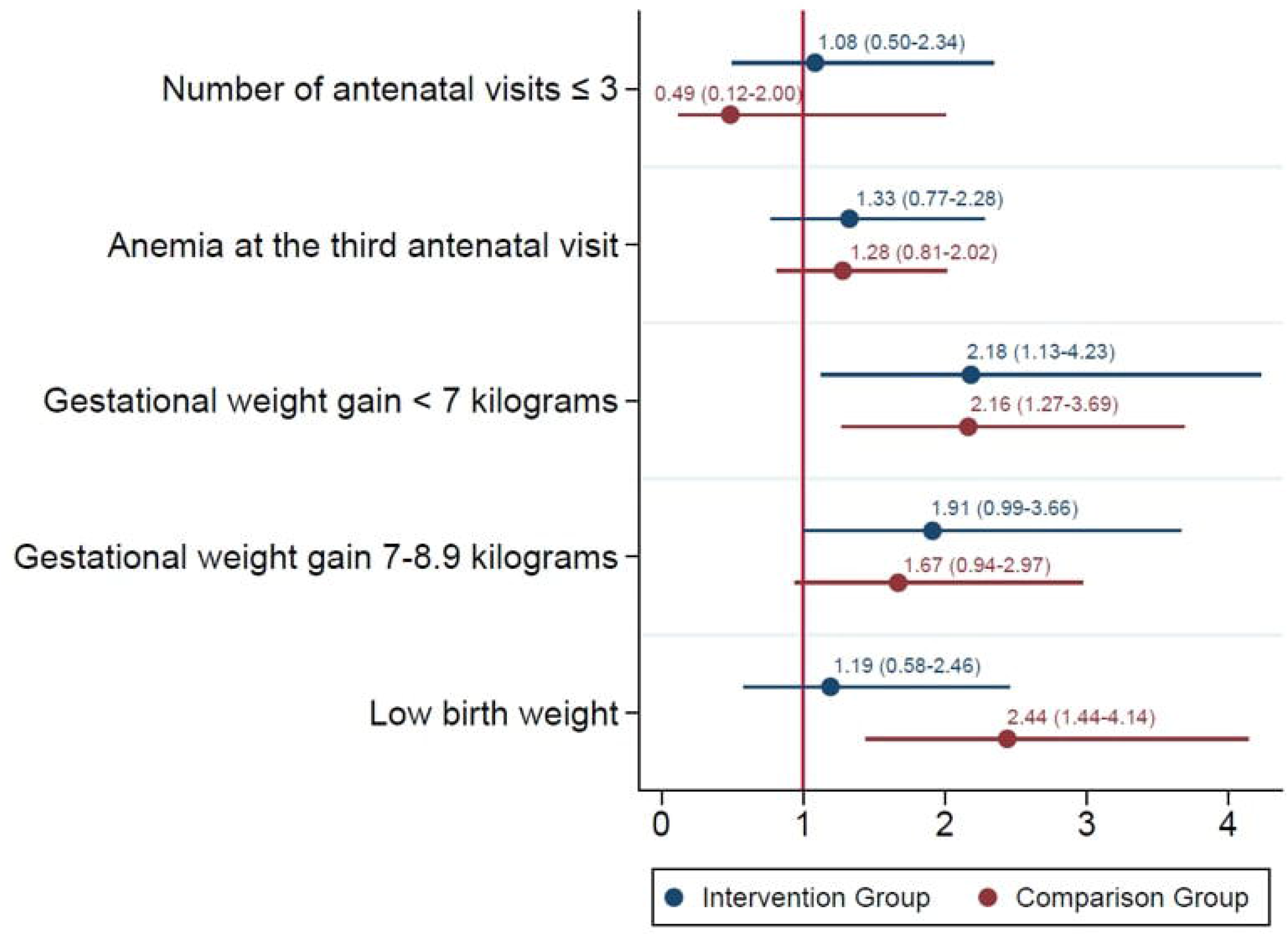
Pregnancy and newborn related factors associated with childhood stunting in the intervention and comparison groups Legend: The odds of stunting amongst children born with low birth weight to women in the comparison group remain statistically significant [OR 2.44 (1.44-4.14)], unlike those amongst children born to women in the intervention group [1.19 (0.58-2.46)].

## Discussion

The findings in this study indicated that a package of antenatal interventions involving systems strengthening approaches and targeted nutritional counselling during pregnancy were associated with improved child nutritional outcomes beyond immediate birth outcomes. The impact of the intervention that was delivered during pregnancy was demonstrable up to 3 years after cessation of the intervention, as children aged between 12-35 months born to women in the intervention group were less likely to be stunted, wasted or underweight in comparison to their counterparts in the non-intervention group.

In this paper, we focus on childhood stunting, as it is a measure of chronic undernutrition and has long lasting repercussions in the individual’s lifecycle. Stunting remains a significant public health concern that is associated with negative outcomes for children and future generations. These associations include increased child morbidity and mortality, increased risk of infections and non-communicable diseases, and poor development and learning capacity(14-16). Stunting is also associated with poor cognitive outcomes, reduced economic productivity, unfavorable maternal reproductive outcomes, as well as higher risk of hypertension, diabetes, and obesity(8).

It is widely recognized that the key ‘window of opportunity’ for reducing stunting is the ∼1000 days from conception until 2 years of age(17). The antenatal period in particular is a crucial window for the expectant mother, since nutritional requirements are high during pregnancy because of the need to support fetal growth. In resource limited settings, this is particularly challenging since women in such settings do not have access to nutrient dense foods. A key component of the intervention provided to pregnant women in this study was to ensure the spot feeding of supplementary nutrition provided to them via the ICDS programme. The take home ration provided to these pregnant women is often shared with family members, thus limiting the extent of nutritional supplementation that the pregnant women actually receive(18). This component of the intervention, along with a strong focus on nutritional counselling of the expectant women in the intervention group seems to have had a lasting effect on childhood nutritional outcomes. The impact of nutrition education and counselling during pregnancy on birth weight and other pregnancy outcomes has been previously evaluated as well. In a meta-analysis of 13 studies, an increase in mean birth weight (+105 g) was noted, but this was significant only when nutrition education/counselling was coupled with nutritional support in the form of food supplements, micronutrient supplements or nutrition safety net interventions(19). The authors, however, did not report the impact of these interventions on birth length. Results from a large trial of multiple micronutrient supplementation in rural Bangladesh (n⍰=⍰28,516 live-born infants) were very similar to the estimates from the meta-analysis: a significant (+54⍰g) effect on mean birth weight, a 12% reduction in low birth weight and a small (+0.2⍰cm) but significant effect on birth length(20). Similarly, a cluster-randomized effectiveness trial (the Rang-Din Nutrition Study) also conducted in Bangladesh found that newborns born to women who received lipid-based nutrient supplements had a reduced risk of stunting (RR: 0.83; 95% CI: 0.71-0.97)(21).

The counselling provided to women in the intervention group in our study also seems to have had a beneficial effect on breastfeeding practices; much larger proportions of women in the intervention group commenced breastfeeding within 1 hour of birth, and exclusively breastfed their children up to 6 months of age. In another study from Tanzania, researchers used a quasi-experimental evaluation design to evaluate the effectiveness of health and nutrition education during the 1000-day window, and found that the proportion of stunted children decreased from 35.9 to 34.2% in intervention sites, and there were increases noted in breast feeding within 1 hour of birth [DID: 7.8%, (95%CI: 2.2–13.4), p = 0.006] and exclusive breast feeding in children under 6 months [DID:20.3%, (95% CI: 10.5–30.1), p = 0.001](22). Although we were not able to assess the postnatal nutritional practices amongst children in both groups in this study, it is likely that the nutritional counselling provided to women in the intervention group had an important impact in the subsequent child feeding practices practiced by the women in the intervention group and may have contributed to the decreased risk of stunting in the children born to these women.

There has been a growing interest in systems strengthening as an approach to improve the implementation of existing interventions to achieve positive maternal and child nutrition outcomes. The Reproductive, Maternal, Newborn, Child, and Adolescent Health (RMNCH+A) strategy in India aims to improve health outcomes across life stages and address inequitable distribution of services for vulnerable groups in poor-performing districts(12). However, performance has been mixed, with variable responsiveness of health systems across states and a lack of robust monitoring and evaluation framework(23). Learnings from the strategy suggest the need for adopting a holistic systems-strengthening based approach towards improving service provision. Systems strengthening has also been identified as a key research priority in India in the COVID-19 pandemic context(24). Our study findings demonstrate the effectiveness of systems strengthening of antenatal care services in improving nutrition outcomes in children. While our findings did not suggest immediate differences between intervention and control groups related to birth weight, an analysis of post-intervention factors associated with stunting showed a higher risk attributable to low birth weight in the comparison compared to the intervention group. This suggests potential sustained impacts of the intervention in the long-term, which could be explained by several biological mechanisms including nutrition during pregnancy and better breastfeeding and child feeding practices. Very few studies have examined long-term impacts of interventions on nutrition outcomes; a recent study showed that India’s mid-day meal school feeding program was associated with intergenerational benefits in child linear growth(25). These findings are of considerable public health significance.

The limitations of this study included a restricted sample size, and a comparison group that was not entirely matched to the intervention group, by virtue of the study design. The follow up of the mother-child dyads in both groups was not on a continuous basis, and we also did not have access to the lengths of the children at birth, and so we are unable to report whether the children were stunted at birth. In addition, we did not have access to data on dietary intake of children or information on nutrition after birth, which is an important determinant for nutrition outcomes including stunting. And lastly, the study was conducted in one state in India, limiting the generalizability of these findings. Despite these limitations, this is amongst the first studies to describe the effect of interventions given during pregnancy and their long-term impact in terms of childhood stunting 2-3 years after the cessation of the intervention.

In conclusion, our study points to the potential of strengthening antenatal care as an effective approach to reduce the prevalence of childhood stunting, the potential impact of focused nutritional counselling to the expectant mother, and its longitudinal effects in terms of improved nutritional outcomes amongst children aged up to 3 years of age. Based on the findings of this study, we would recommend incorporating baseline maternal BMI measurements and monthly eight monitoring into routine antenatal care, incorporating systems strengthening measures to ensure the effective uptake of maternal nutritional supplementation during the antenatal period, and providing targeted dietary counselling based on the BMI and overall nutritional status of the expectant mother. These simple measures provided during the antenatal period would go a long way in not only improving maternal outcomes, but also can have distal beneficial effects on childhood stunting beyond the immediate post-natal period.

## Data Availability

All data produced in the present study are available upon reasonable request to the authors

## Abbreviations

BMI: Body Mass Index
ICDS: Integrated Child Development Services
NFHS: National Family Health Survey
RMNCH+A: Reproductive, Maternal, Newborn, Child, and Adolescent Health

## Figure titles and legends

Supplementary figure 1: Impact of the intervention on nutritional status in children aged between 12-23 months

Legend: Note: *p<0.05, **p<0.01, ***p<0.001 denotes differences in RR between intervention and comparison groups

Supplementary figure 2: Impact of the intervention on nutritional status in children aged between 24-35 months

Legend: Note: *p<0.05, **p<0.01, ***p<0.001 denotes differences in RR between intervention and comparison groups

## References

1. World Health Organization. (2014). Global nutrition targets 2025: policy brief series. Available at https://www.who.int/publications/i/item/9789241549912. Accessed October 14, 2022.

2. United Nations. (2015). Sustainable Development Goals. Available at https://www.undp.org/sustainable-development-goals. Accessed October 14, 2022.

3. World Health Organization. Levels and trends in child malnutrition. UNICEF / WHO / World Bank Group Joint Child Malnutrition Estimates. Key findings of the 2021 edition. Available at https://www.undp.org/sustainable-development-goals. Accessed October 14, 2022.

4. International Institute for Population Sciences (IIPS) and ICF. 2020. National Family Health Survey (NFHS)-5. Available at https://dhsprogram.com/pubs/pdf/OF43/NFHS-5_India_and_State_Factsheet_Compendium_Phase-II.pdf. Accessed October 14, 2022.

5. International Institute for Population Sciences - IIPS/India, ICF. India National Family Health Survey NFHS-4 2015-16. Mumbai, India: IIPS and ICF; 2017. Available at https://dhsprogram.com/publications/publication-fr339-dhs-final-reports.cfm. Accessed October 14, 2022.

6. Black RE, Victora CG, Walker SP, Bhutta ZA, Christian P, de Onis M, et al. Maternal and child undernutrition and overweight in low-income and middle-income countries. Lancet (London, England). 2013;382(9890):427–51.

7. Grantham-McGregor S, Cheung YB, Cueto S, Glewwe P, Richter L, Strupp B. Developmental potential in the first 5 years for children in developing countries. Lancet (London, England). 2007;369(9555):60–70.

8. Dewey KG, Begum K. Long-term consequences of stunting in early life. Matern Child Nutr. 2011;7 Suppl 3:5–18.

9. Lowensohn RI, Stadler DD, Naze C. Current Concepts of Maternal Nutrition. Obstet Gynecol Surv. 2016;71(7):413–26.

10. Bhutta ZA, Das JK, Rizvi A, Gaffey MF, Walker N, Horton S, et al. Evidence-based interventions for improvement of maternal and child nutrition: what can be done and at what cost? Lancet (London, England). 2013;382(9890):452–77.

11. Mukherjee SG, Sen P, Shah NN. Ensuring pregnancy weight gain: An integrated community-based approach to tackle maternal nutrition in India. Field Exchange issue 61, November 2019.

12. Ministry of Health & Family Welfare, Government of India. A Strategic Approach to Reporoductive, Maternal, Newborn, Child and Adolescent Health. (RMNCH+A) in India. Available at https://nhm.gov.in/images/pdf/RMNCH+A/RMNCH+A_Strategy.pdf. Accessed October 14, 2022.

13. World Health Organization. WHO recommendations on antenatal care for a poitive pregnancy experience. Available at https://www.who.int/publications/i/item/9789241549912. Accessed ctober 14, 2022.

14. Victora CG, Christian P, Vidaletti LP, Gatica-Domínguez G, Menon P, Black RE. Revisiting maternal and child undernutrition in low-income and middle-income countries: variable progress towards an unfinished agenda. The Lancet. 2021;397(10282):1388–99.

15. De Sanctis V, Soliman A, Alaaraj N, Ahmed S, Alyafei F, Hamed N. Early and Long-term Consequences of Nutritional Stunting: From Childhood to Adulthood. Acta Biomed. 2021;92(1):e2021168.

16. de Onis M, Branca F. Childhood stunting: a global perspective. Maternal & child nutrition. 2016;12 Suppl 1(Suppl 1):12-26.

17. Victora CG, de Onis M, Hallal PC, Blössner M, Shrimpton R. Worldwide timing of growth faltering: revisiting implications for interventions. Pediatrics. 2010;125(3):e473–80.

18. Talati KN, Nimbalkar S, Phatak A, Patel D. Take home ration in ICDS programmes: Opportunities for integration with health system for improved utilisation via Mamta card and eMamta. BMJ Glob Health. 2016;1:A7–A8.

19. Girard AW, Olude O. Nutrition education and counselling provided during pregnancy: effects on maternal, neonatal and child health outcomes. Paediatric and perinatal epidemiology. 2012;26 Suppl 1:191–204.

20. West KP, Jr., Shamim AA, Mehra S, Labrique AB, Ali H, Shaikh S, et al. Effect of maternal multiple micronutrient vs iron-folic acid supplementation on infant mortality and adverse birth outcomes in rural Bangladesh: the JiVitA-3 randomized trial. JAMA. 2014;312(24):2649–58.

21. Mridha MK, Matias SL, Chaparro CM, Paul RR, Hussain S, Vosti SA, et al. Lipid-based nutrient supplements for pregnant women reduce newborn stunting in a cluster-randomized controlled effectiveness trial in Bangladesh. Am J Clin Nutr. 2016;103(1):236–49.

22. Elisaria E, Mrema J, Bogale T, Segafredo G, Festo C. Effectiveness of integrated nutrition interventions on childhood stunting: a quasi-experimental evaluation design. BMC Nutr. 2021;7(1):17.

23. Taneja G, Sridhar VS, Mohanty JS, Joshi A, Bhushan P, Jain M, et al. India’s RMNCH+A Strategy: approach, learnings and limitations. BMJ Glob Health. 2019;4(3):e001162.

24. Mehta K, Zodpey S, Banerjee P, Pocius SL, Dhaliwal BK, DeLuca A, et al. Shifting research priorities in maternal and child health in the COVID-19 pandemic era in India: A renewed focus on systems strengthening. PLoS One. 2021;16(8):e0256099.

25. Chakrabarti S, Scott SP, Alderman H, Menon P, Gilligan DO. Intergenerational nutrition benefits of India’s national school feeding program. Nat Commun. 2021;12(1):4248.

