## Supplementary figures and images for "Systems strengthening approach during antenatal care improves maternal nutrition and reduces childhood stunting in West Bengal, India"

### Supplementary fig 1

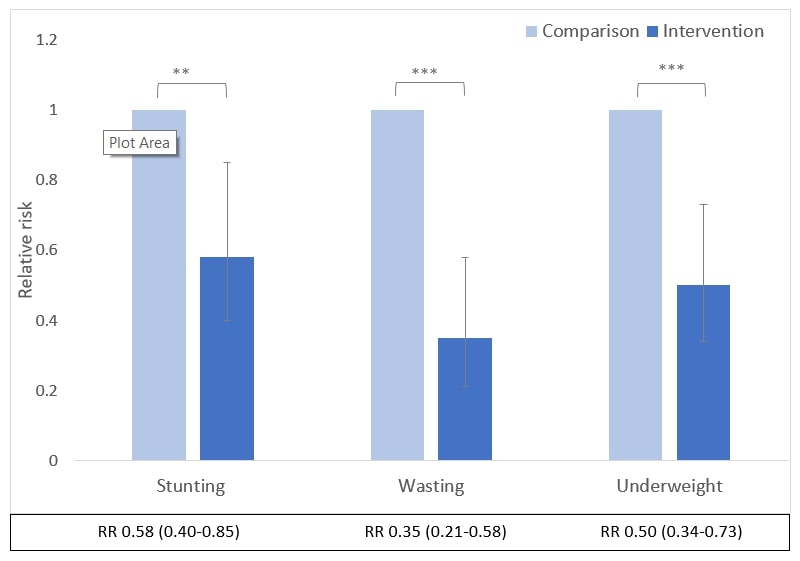

### Supplementary fig. 2

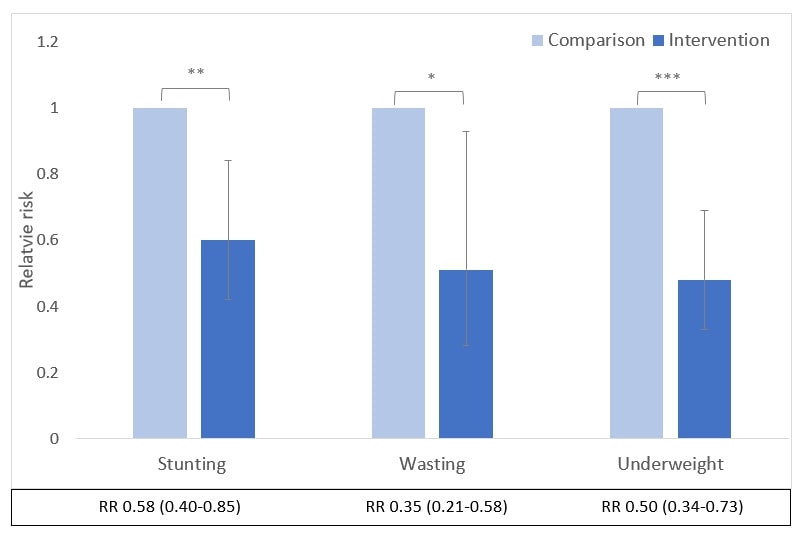
